# Simulated Diagnostic Performance of Ultra-Low-Field MRI: Harnessing Open-Access Datasets to Evaluate Novel Devices

**DOI:** 10.1101/2021.07.02.21259789

**Authors:** T. Campbell Arnold, Steven N. Baldassano, Brian Litt, Joel M. Stein

## Abstract

The purpose of this study is to demonstrate a method for virtually evaluating novel imaging devices using machine learning and open-access datasets, here applied to a new, ultra-low-field strength (ULF), 64mT, portable MRI device. Paired 3T and 64mT brain images were used to develop and validate a transformation converting standard clinical images to ULF-quality images. Separately, 3T images were aggregated from open-source databases spanning four neuropathologies: low-grade glioma (LGG, N=76), high-grade glioma (HGG, N=259), stroke (N=28), and multiple sclerosis (MS, N=20). The transformation method was then applied to the open-source data to generate simulated ULF images for each pathology. Convolutional neural networks (DenseNet-121) were trained to detect pathology in axial slices from either 3T or simulated 64 mT images, and their relative performance was compared to characterize the potential diagnostic capabilities of ULF imaging. Algorithm performance was measured using area under the receiver operating characteristic curve. Across all cohorts, pathology detection was similar between 3T and simulated 64mT images (LGG: 0.97 vs. 0.98; HGG: 0.96 vs. 0.95; stroke: 0.94 vs. 0.94; MS: 0.90 vs 0.87). Pathology detection was further characterized as a function of lesion size, intensity, and contrast. Simulated images showed decreasing sensitivity for lesions smaller than 4 cm^2^ (∼2.25 cm in diameter). While simulations cannot replace prospective trials during the evaluation of medical devices, they can provide guidance and justification for prospective studies. Simulated data derived from open-source imaging databases may facilitate testing and validation of new imaging devices.

**Highlights:**

- Ultra-low-field, point-of-care MRI has potential to detect a range of pathologies including brain tumors, strokes, and multiple sclerosis. However, determining the diagnostic capabilities and appropriate use case for such devices requires further prospective studies.
- Open-source image datasets provide a powerful tool for accelerating imaging research and enable simulated trials that can guide prospective clinical trials or device development.

## 1. Introduction

Modern medical imaging has become a mainstay of patient care, particularly in the diagnosis and management of patients with neurologic disease. While the availability of imaging technology has dramatically increased worldwide in recent decades, the expense and operational complexity of standard imaging systems limits access in underserved areas and developing countries [1]. This so-called “radiology divide” leaves about ninety percent of the world’s population without access to magnetic resonance imaging (MRI) [2] and almost two-thirds of the population without even basic imaging technology such as ultrasound and X-ray radiography [3– 5].

Ultra-low-field strength (ULF, <0.1T) MRI systems aim to make MRI more accessible, promising lower cost, portability, fewer magnetic field-related safety concerns, and ease of use [6]. Such devices could decrease healthcare expenditures, improve availability in underserved areas, and provide a convenient and ionizing-radiation-free modality for routine or monitoring studies. ULF MRI systems may be suitable for hospitalized patients, such as those in intensive care units or isolation wards, for whom transport to a standard clinical scanner carries unacceptable risk [7–9]. More broadly, ULF MRI units could potentially be used in ambulances, emergency departments, physician’s offices or rural clinics [10,11].

While ULF MRI presents clear practical advantages, these systems produce images with lower signal-to-noise than their high-field strength (HF) counterparts and are largely designed to complement, and not replace, standard MRI. Prior to deployment for clinical use, the diagnostic capabilities of novel imaging technologies such as ULF MRI should be evaluated across a wide range of patients and pathology. The standard approach for device evaluation, improvement, and optimization requires recruiting large numbers of patients and manual image review by radiologists. This process is costly and time-consuming, which can limit the device development cycle. Moreover, selecting target use cases is difficult without basic information about device sensitivity. A complementary approach is to simulate ULF images from existing HF data, leveraging large publicly accessible HF imaging databases. Such databases, typically compiled for machine learning competitions [12–15] or collaborative research programs [16,17], span broad ranges of pathology and offer a wealth of information for retrospective analysis [18].

Here, we propose a generalizable approach to use open-access imaging databases to evaluate novel imaging devices. We employ an empiric method to transform existing open-access images to a custom domain. We use this simple method to convert high-resolution 3T MR images aggregated from several open-access databases to lower-quality images designed to mimic images acquired on an ULF MRI scanner. Separate convolutional neural networks are then trained to detect pathology in axial slices from the HF or simulated ULF images, and detection performance is compared between image pairs to characterize the potential diagnostic capabilities of ULF MRI. While automated lesion detection in simulated images does not guarantee detection on the actual device, simulated performance may help indicate whether pursuing a prospective study for a given application is promising. Applied here to ULF MRI, this simulated trial approach offers a broadly applicable means for evaluating and optimizing novel medical imaging technologies to complement traditional clinical trials.

## 2. Materials & Methods

### 2.1. Data collection

We collected data from six adult patients with known or suspected hydrocephalus imaged same-day on a 64mT ULF MRI scanner (Hyperfine) and a standard 3T HF MRI system (Siemens). Data was collected as part of an ongoing research study approved by the University of Pennsylvania Institutional Review Board and patients provided informed consent. Axial fluid-attenuated inversion recovery (FLAIR) images covering the whole brain were collected on each scanner. Image resolution was 1.5 x 1.5 x 5 mm for ULF images and ranged from isotropic to 0.78 x 0.78 x 3 mm for HF images.

Separately, retrospective axial FLAIR images obtained at 3T for a range of pathologies were aggregated from several open-access sources [12–15,19]. Pathologies consisted of high-grade glioma (HGG, N=259), low-grade glioma (LGG, N=76), stroke (N=28), and multiple sclerosis (MS, N=20) [12–15]. Each dataset contained manual segmentations of lesions, which generally manifest as hyperintense areas on FLAIR imaging. These datasets incorporate a range of lesion sizes and signal intensities that can be quantified using the provided lesion segmentations. Note that HGG and LGG lesion segmentations on FLAIR imaging include areas that may represent vasogenic edema or non-enhancing infiltrative neoplasm as well as enhancing components when present. Additionally, non-lesional control subjects (N=5) were drawn from the OASIS3 dataset [19]. **Table 1** contains information about pathology datasets used in this retrospective study and related web addresses as of publication are listed in section *2.8 Code and data availability*.

**Table 1.** List of open-access neuropathology datasets used in this study. Abbreviations: High Grade Glioma (HGG), Low Grade Glioma (LGG), Multiple Sclerosis (MS).

| Dataset | BraTS 2019 | ISLES 2015 | MICCAI 2008 | MS-SEG-2016 |
| --- | --- | --- | --- | --- |
| Pathology | Glioma (HGG & LGG) | Stroke | Multiple Sclerosis (MS) | Multiple Sclerosis (MS) |
| Subjects | 335 (259 HGG, 76 LGG) | 28 | 10 | 10 |
| Center/Scanners | 19 | 2 | 2 | 3 |
| Classes | 3 (enhanced, non-enhanced, edema) | 1 (infarct) | 1 (MS lesion) | 1 (MS lesion) |

### 2.2. Simulated image generation: High-field to ultra-low-field MRI

To generate simulated ULF images from HF data, we employed a simple image transformation using 3T and 64mT image pairs from three subjects. Transformation steps are listed in **Fig. 1A** and include registration, brain extraction, re-slicing, Gaussian smoothing, and noise filtering. To match ULF image quality, smoothing and noise filters were parameterized and differences in histogram features between real and simulated images were minimized. The objective function was comprised of the first three statistical moments (mean, standard deviation, and skewness). An example HF/ULF pair and the simulated ULF image can be seen in **Fig. 1B** with intensity histograms shown in **Fig. 1C**. The transformation was validated using data from three additional subjects. The image transformation method was quantitatively assessed using the gradient entropy, *F*, as a measure of perceived diagnostic image quality. This metric was derived by McGee et al. [20]:

**Fig. 1.**
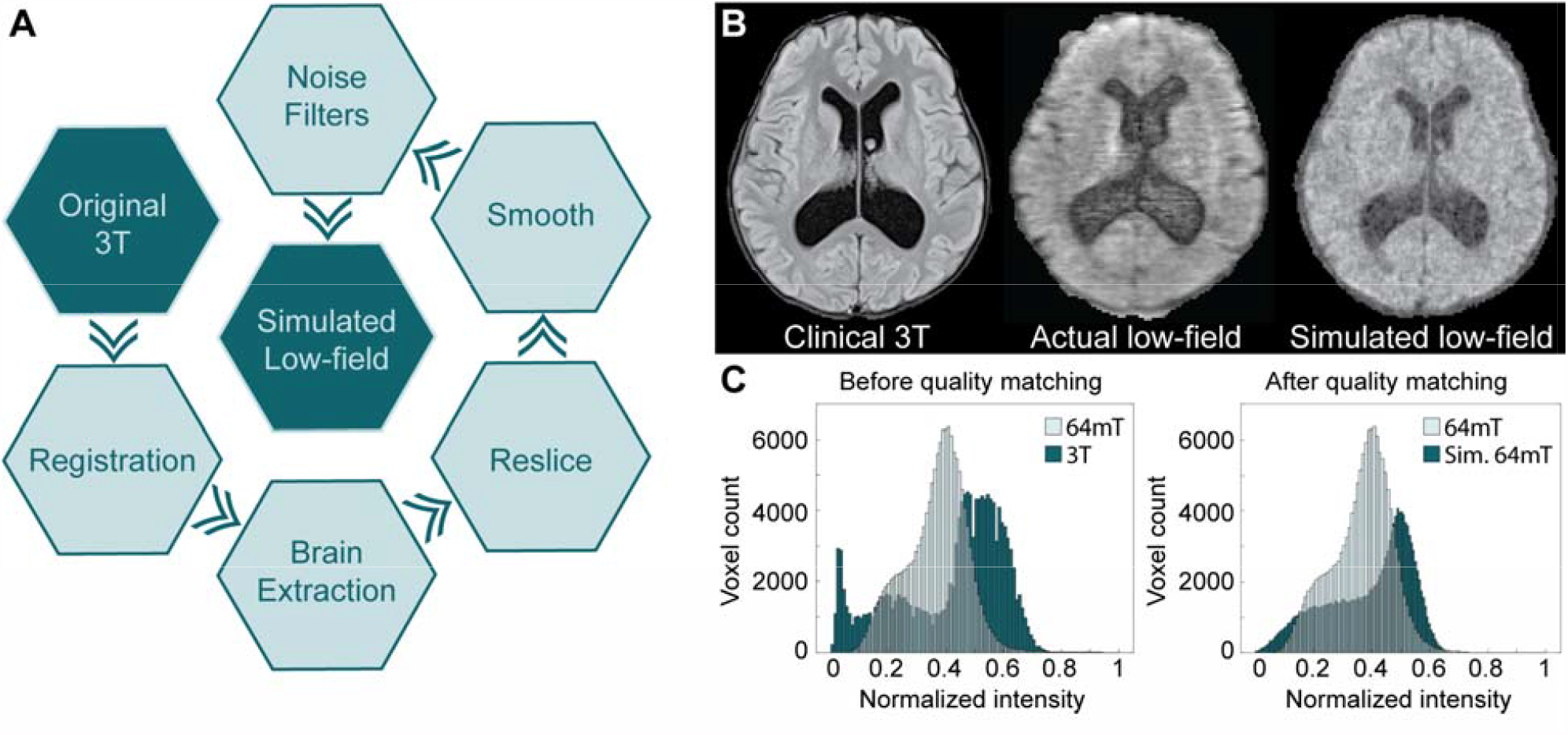
Generating simulated ultra-low-field (ULF) images. **(A)** Steps in the image processing pipeline from the original 3T image (upper-left) to the simulated ULF version (center). **(B)** Example skull-stripped axial FLAR images from a clinical 3T (left) and a 64mT ULF MRI scanner (center). The 3T image was passed through the image transformation pipeline to produce the simulated ULF image (right). **(C)** To generate simulated ULF images resembling actual ULF images, histogram features (mean, standard deviation, and skewness) were used to guide image transformation. The intensity histogram distributions relative to actual 64 mT images are shown before (left) and after (right) transformation.

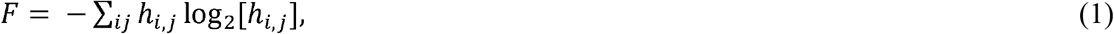

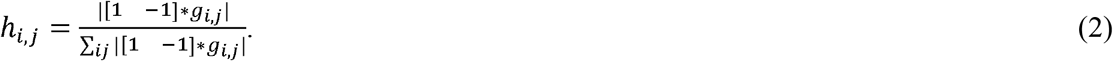

where *g*_*i,j*_ is the pixel value at coordinate *i,j* and * represents the convolution operation. Of 24 metrics evaluated, McGee et al. found gradient entropy (Equation 1) to be the most strongly correlated with perceived image quality when applied to structural MRI. The transformation was applied to the HF images collected from open-access datasets to produce simulated ULF images for glioma, stroke, and multiple sclerosis.

### 2.3. Modulating lesion contrast

To assess the relationship between lesion contrast and detection accuracy, we prepared additional simulated ULF images from the HGG dataset by decreasing signal intensity within the segmented lesions. Lesion intensity values were scaled independently from surrounding brain tissue in 20% increments over a range from 100% (original intensity) to 0% (isointense with background tissue). Isointensity was defined as mean lesion intensity equal to mean intensity of non-lesional tissue in the same slice. As described subsequently, separate classifiers were trained to identify lesions at each intensity level, allowing for the decoupling of intensity contrast and structural abnormalities in classifier performance. Note that because of structural abnormalities caused by large tumors, such as mass effect, midline shift, and ventricular effacement, as well as intensity heterogeneity within lesions, even isointense lesions may retain some structural and signal alterations after contrast modulation. To minimize the effect of intensity heterogeneity, we restricted our analysis to subjects within the top 50% of lesion homogeneity (N=130), as defined by within tumor signal to noise ratio (SNR). Accurate detection of isointense lesions should therefore be primarily driven by structural abnormalities.

### 2.4. Model Architecture

A convolutional neural network model was used to identify pathology in each high-field and simulated ultra-low-field dataset. Model construction and training was carried out using the Keras API [21] with TensorFlow [22,23] backend. Model architecture consisted of the DenseNet-121 network [24,25] with initial weights pre-trained on the ImageNet database [26] and four additional densely-connected layers using Xavier initialization. This architecture was consistent across datasets (Supplemental Fig. 1).

### 2.5. Model Training

For each dataset, a unique model was trained to perform binary classification of axial slices (lesion present vs. lesion absent). Separate models were trained on the HF and simulated ULF images. Slices were labeled as “lesion present” if at least one pixel from the ground-truth lesion segmentations was present. Each dataset was divided (∼9:1 split) into “training” and “test” datasets (Supplemental Table 1). Each subject was confined to either the training or test dataset. Training image order was randomly shuffled. All reported performance metrics are derived from held-out test data. Models were trained for 100 epochs using the Nadam optimizer, a learning rate of 0.002 with decay [27], and a batch size of 32. Batch size was chosen to accommodate VRAM of Titan X GPU. Training data were augmented using random horizontal flipping. Training parameters were consistent across datasets.

### 2.6. Model Evaluation

Classification performance was evaluated using two metrics: (1) area under curve (AUC) of the receiver operating characteristic (ROC) and (2) F1 score (harmonic mean of precision and recall). A random chance null model (performance averaged over 1000 trials) was included for comparison.

Class activation maps (CAMs) were generated from shallow and deep convolutional layers to identify discriminative image regions [28]. CAMs were constructed using the output feature map of a given convolutional layer with each feature map channel weighted by the lesion class gradient. Conceptually, CAMs help to interpret model function by visualizing image areas that are driving the model’s classification decision.

In addition to slice-by-slice classification performance, models were evaluated on a per-subject basis. For each test subject, the models assigned a classification score to all axial slices. A sliding convolutional filter was used to determine the mean classification score over several adjacent slices (approximately 1.5 cm in the z-axis) (Supplemental Fig. 2).

**Fig. 2.**
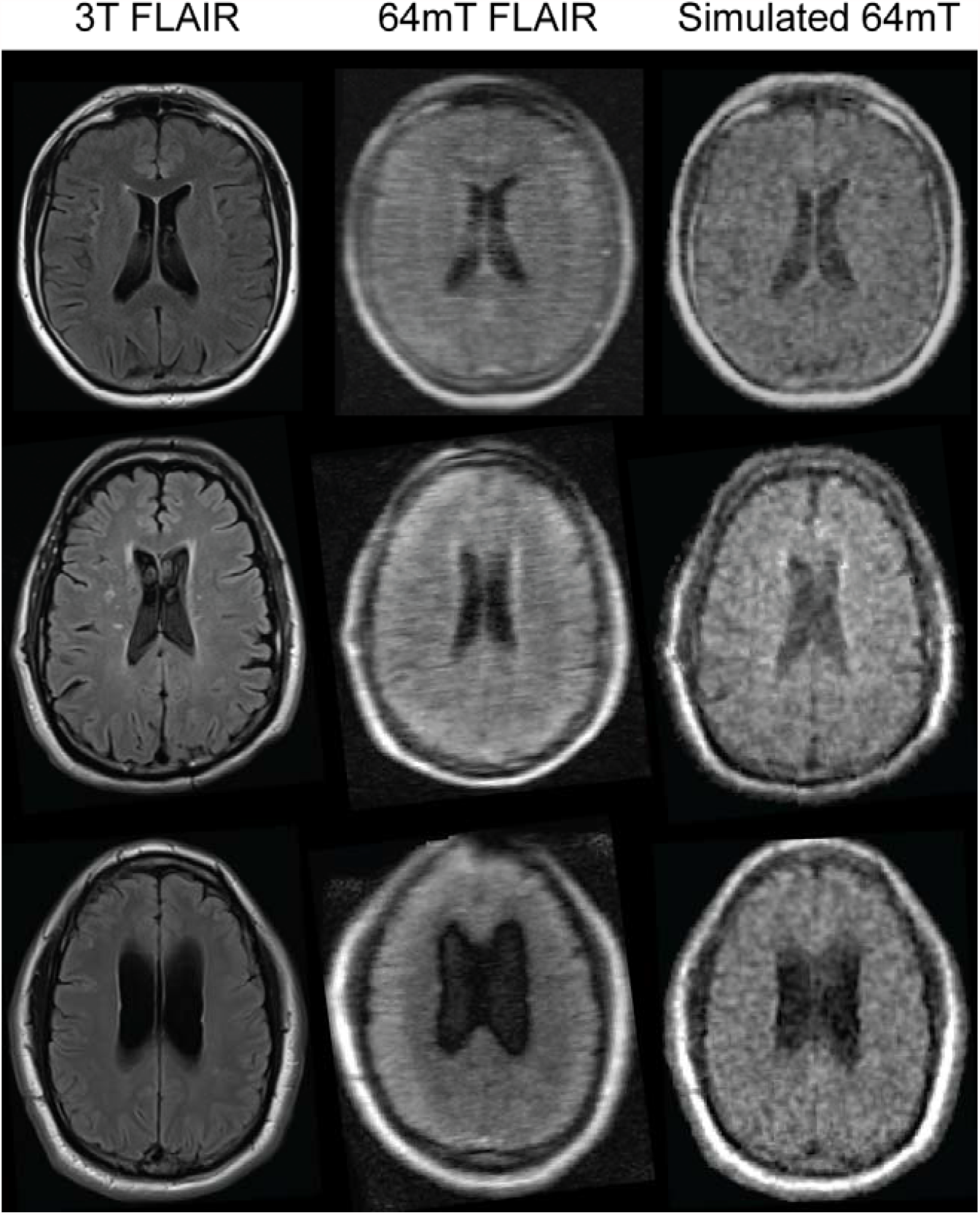
Validation of image transformation method. 3T images (column 1), simulated 64mT images (column 2), and actual 64mT images (column 3) are shown for three additional subjects. These subjects were not included in the training set used to develop the automated transformation.

### 2.7. Statistical Analysis

ROC curves were compared using DeLong’s test, implemented using the pROC R package [29]. Significance of logistic regression parameters was determined by Wald test.

### 2.8. Code and data availability

All code related to simulated image generation, classifier design, and statistical analysis can be found at: https://github.com/penn-cnt/Arnold_simulated_clinical_trial. We are grateful to the researchers that published the well-curated, publicly available datasets used in this study. As of publication, these data can be found at MS-SEG 2008 [12]: http://www.ia.unc.edu/MSseg/, BraTS 2019 [13]: https://www.med.upenn.edu/cbica/brats2019.html, MS-SEG 2016 [14]: https://portal.fli-iam.irisa.fr/english-msseg/, ISLES 2015 [15]: http://www.isles-challenge.org/ISLES2015/, OASIS3 [19]: https://www.oasis-brains.org/.

## 3. Results

### 3.1. Image Transformation Validation

The image transformation method was validated on data from three additional subjects not included during the transformation fitting step. For each subject, a standard 3T FLAIR image was transformed into a simulated ULF image for comparison against the authentic ULF ground truth. Representative images from each subject are shown in **Fig. 2**.

Perceived diagnostic image quality was quantitatively assessed using the entropy of the MR image gradient. Lower gradient entropy indicates sharper edges and correlates strongly with higher perceived image quality on MRI [20]. Gradient entropy of HF images (mean ± standard deviation: 5.91± 0.54) was significantly lower than both the real ULF images (7.89± 0.90) and simulated ULF (7.42± 0.73) images (p<0.0001, t-test). While there was a statistical difference between the gradient entropy of real and simulated ULF images (p<0.05), the effect size was dramatically reduced compared to the original HF images (0.47 vs 1.98). While perceived quality was modestly higher in simulated ULF images, there was substantial overlap of gradient entropy with real ULF images, indicating similar image quality between simulated and real images.

### 3.2. Comparing pathology detection in standard and simulated ultra-low-field images

Deep learning models were trained to perform binary classification of images in each disease cohort using either HF or simulated ULF MRI. ROC curves for each cohort are shown in **Fig. 3**. Despite significant image degradation, per-slice classifier performance was similar for HF and simulated ULF MRI across all pathologies (**Table 2**). As expected, accuracy on subtle pathology (MS lesions) was lower than more prominent pathology for both HF and simulated ULF MRI datasets. The performance achieved is comparable to previous benchmarks using deep learning for detection of brain masses [30,31], and MS lesions [32], and significantly exceeded null models in all cohorts tested.

**Table 2.** Performance metrics for each pathology type using standard or simulated low-field images. Abbreviations: High Grade Glioma (HGG), Low Grade Glioma (LGG), Multiple Sclerosis (MS), Area Under the Curve (AUC).

| Pathology | Standard AUC | Low-Field AUC | p<sub>AUC</sub> | Standard F1 | Low-Field F1 | Null Model F1 |
| --- | --- | --- | --- | --- | --- | --- |
| HGG | 0.972 | 0.978 | 0.16 | 0.920 | 0.896 | 0.481±0.01 |
| LGG | 0.957 | 0.949 | 0.61 | 0.885 | 0.880 | 0.468±0.02 |
| Stroke | 0.936 | 0.940 | 0.83 | 0.772 | 0.761 | 0.485±0.02 |
| MS | 0.896 | 0.873 | 0.49 | 0.766 | 0.745 | 0.442±0.02 |

**Fig. 3.**
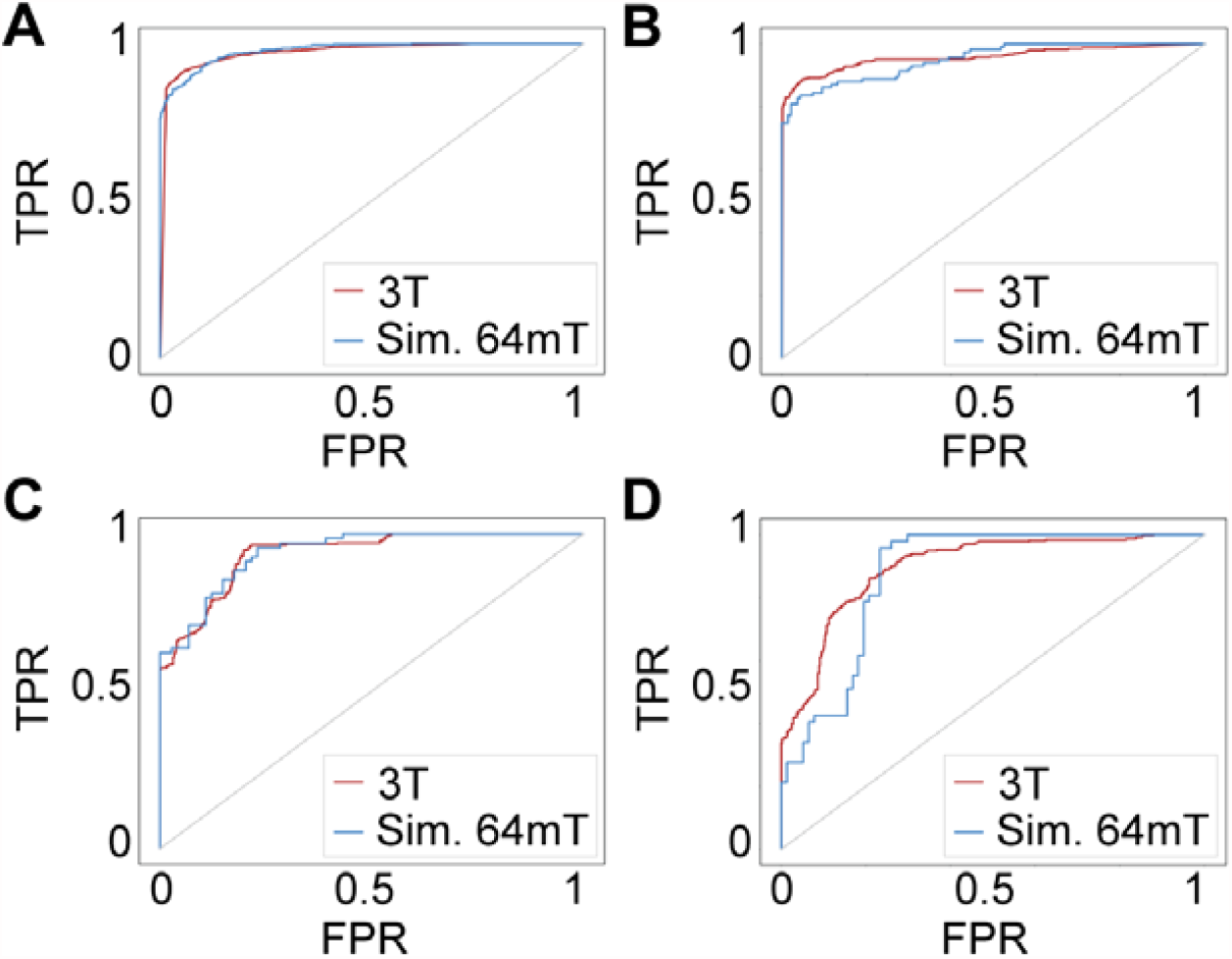
Pathology detection performance. Receiver operating characteristic (ROC) curves shown for binary classification of images with and without pathology for high-grade glioma **(A)**, low-grade glioma **(B)**, ischemic stroke **(C)**, and multiple sclerosis **(D)**. Curves are shown for standard (3T) and simulated low-field images. Abbreviations: True positive rate (TPR), False positive rate (FPR), Simulated (Sim.).

### 3.3. Characterizing ultra-low-field pathology detection

To broadly characterize pathology detection capabilities in simulated ULF images, detection sensitivity was aggregated across all pathologies and modeled using a logistic regression as a function of lesion size and intensity as shown in **Fig. 4A & 4B**. For both HF and simulated ULF images, sensitivity was more strongly associated with lesion size, though both parameters reached statistical significance (standard: z_size_=13.5, p_size_<2e-16, z_intensity_=9.2, p_intensity_<2e-16; simulated ultra-low-field: z_size_=8.5, p_size_<2e-16, z_intensity_=3.9, p_intensity_<9e-5). HF imaging outperformed simulated ULF imaging for detection of smaller or less intense lesions as shown in **Fig. 4C**. While performance did not vary significantly between HF and simulated ULF images across the cohorts as a whole, sensitivity differences in this subgroup analysis suggests a performance drop-off when using ULF imaging for 1-4 cm^2^ lesions (**Fig. 4A & 4B**).) These findings are agnostic to pathology type and may serve as generalizable performance guidelines for FLAIR at 64mT in yet-untested patient populations.

**Fig. 4.**
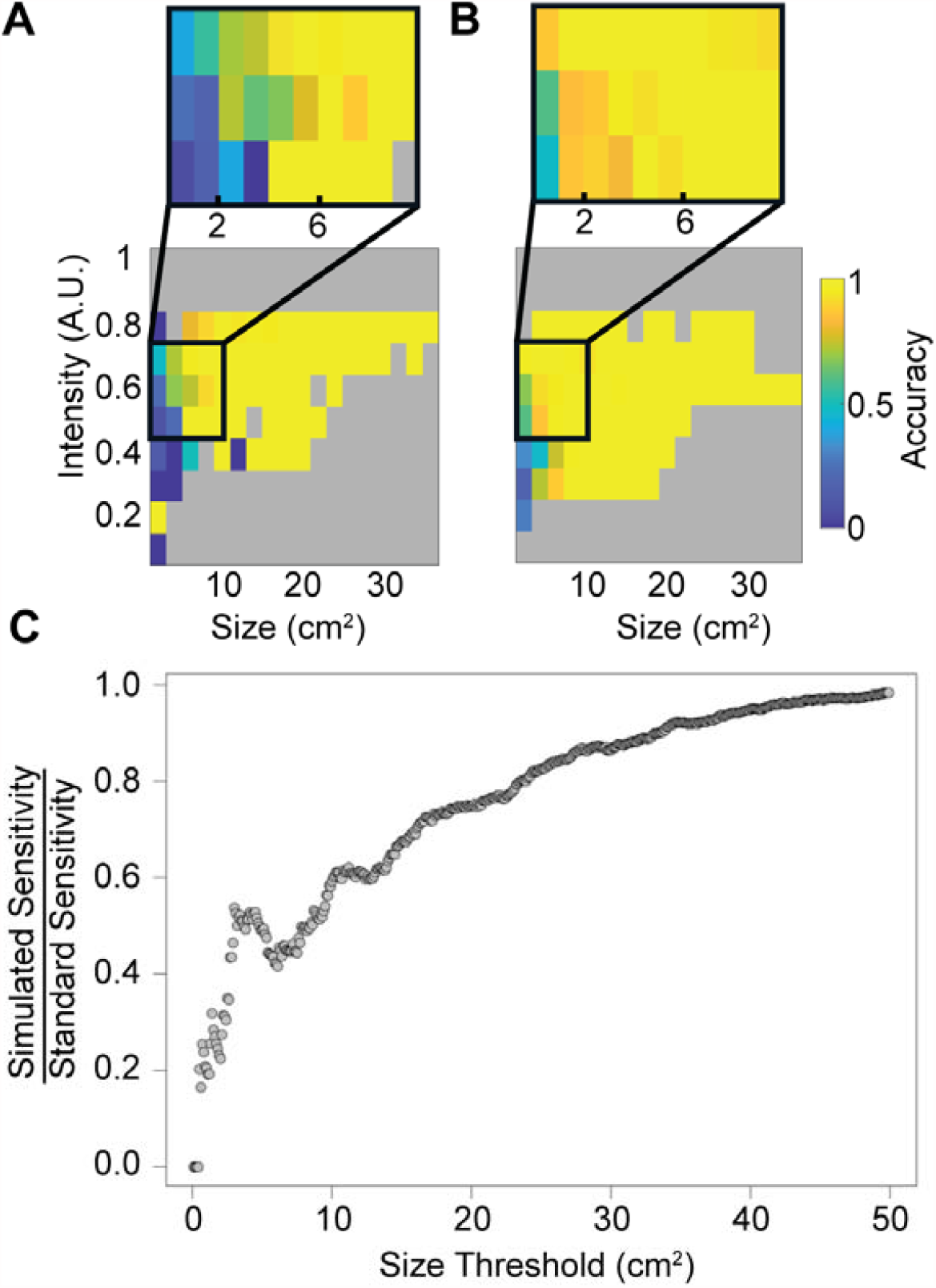
Detection sensitivity as a function of lesion size and scaled intensity. Sensitivity of the deep learning classifiers for detecting lesions in the validation set is shown in the **(A)** simulated ULF and **(B)** standard HF images. Areas highlighting discrepancies between the datasets are highlighted in image insets. **(C)** Sensitivity of lesion detection in simulated ULF images relative to HF images. Each point represents the sensitivity ratio measured on all lesions smaller than the given size threshold. Note that sensitivity is similar between image types when averaged over all lesions but differs significantly when restricted to smaller lesions.

### 3.4. Patient-level classification

In addition to per-slice performance, we assessed pathology detection on a per-subject basis. Algorithms were evaluated over 17 held-out subjects (three MS, four stroke, five HGG, five LGG) and achieved 100% sensitivity in both HF and simulated ULF images. The LGG classifier was also evaluated using five control subjects, and correctly identified all subjects as non-lesional (100% specificity in both HF and simulated ULF images) as shown in **Fig. 5A & 5B**.

**Fig. 5.**
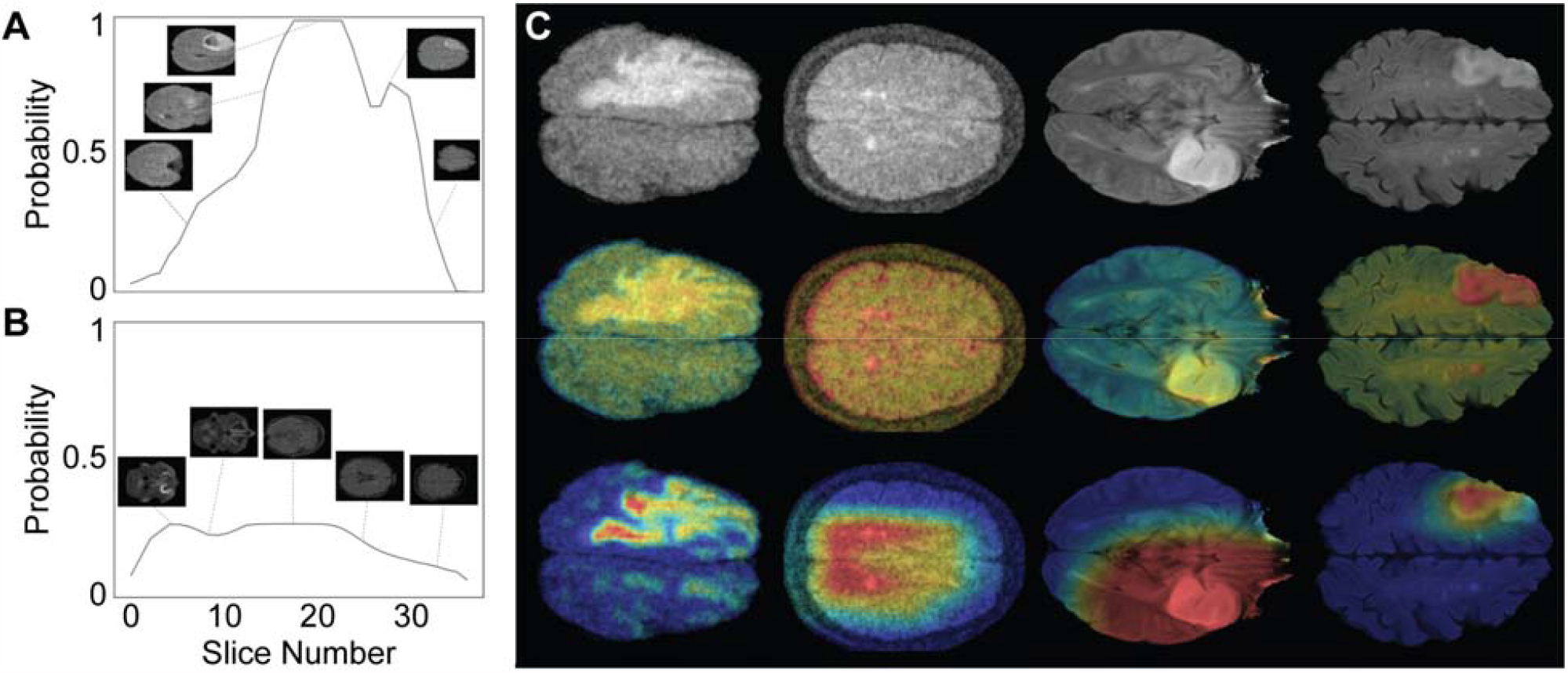
Model validation and interpretability. Panel A and B provide examples of per-subject pathology detection. Convolutional filters were used to generate average lesion probability values across several adjacent axial slices. A threshold value for subject-level classification was determined empirically by maximizing per-subject classification accuracy in the training set. Sample plots are shown for a subject with HGG **(A)** and a control patient **(B)** using simulated low-field imaging. **(C)** Class activation mapping. Row 1: Sample images of high-grade glioma (low-field), multiple sclerosis (low-field), low-grade glioma (standard), and ischemic stroke (standard). Row 2: CAMs generated from shallow network layer for each pathology. Row 3: CAMs generated from deepest convolutional network layer for each pathology.

### 3.5. Class activation mapping

We used class activation mapping to probe which image regions were driving algorithm decisions as shown in **Fig. 5C**. As expected, areas containing pathology are the primary drivers of classification at both shallow and deep layers. The deep CAM for the MS model also reveals that this model attends to periventricular areas known to be clinically important for lesion identification. These findings are reassuring that the tested models are detecting pathology of interest as expected and may have empirically captured features of the typical disease distribution. It is important to note that CAMs serve as an approximate representation of model attention, and each convolutional model layer has a unique CAM. While interpretation of CAMs alone is difficult due to the nonlinear nature of neural networks, the CAMs and the subject level visualizations provide convergent evidence that our models are attending to pathological features.

### 3.6. Determining the effect of lesion intensity on detection

A potential limitation of the simulated image generation method is that it does not account for possible changes in ULF lesion to background tissue contrast relationships due to differences in relaxation times or pulse sequences. Here, we quantify detection robustness by measuring performance over a range of lesion contrasts (outlined in *2*.*3. Modulating lesion contrast*) for HGG images. This approach also allows us to assess the relative importance of lesion to background tissue contrast in comparison to structural distortion from large brain tumors.

ROC curves for detection of HGGs are shown in **Fig. 6**. Compared to detection of full-intensity tumors (AUC=0.972), there was a statistically significant decrease in performance for tumors with relative contrast of 60% or less (AUC_80_=0.972, p=NS; AUC_60_=0.943, p=0.01; AUC_40_=0.905, p=2.0e-5; AUC_20_=0.901, p=2.2e-6; AUC_0_=0.874, p=1.7e-8). While contrast has a substantial impact on lesion detection, an AUC of 0.874 and F1 of 0.71 is achieved even for isointense lesions, indicating that in this dataset even with reduced lesion contrast large pathology could be identified due to structural deformation.

**Fig. 6.**
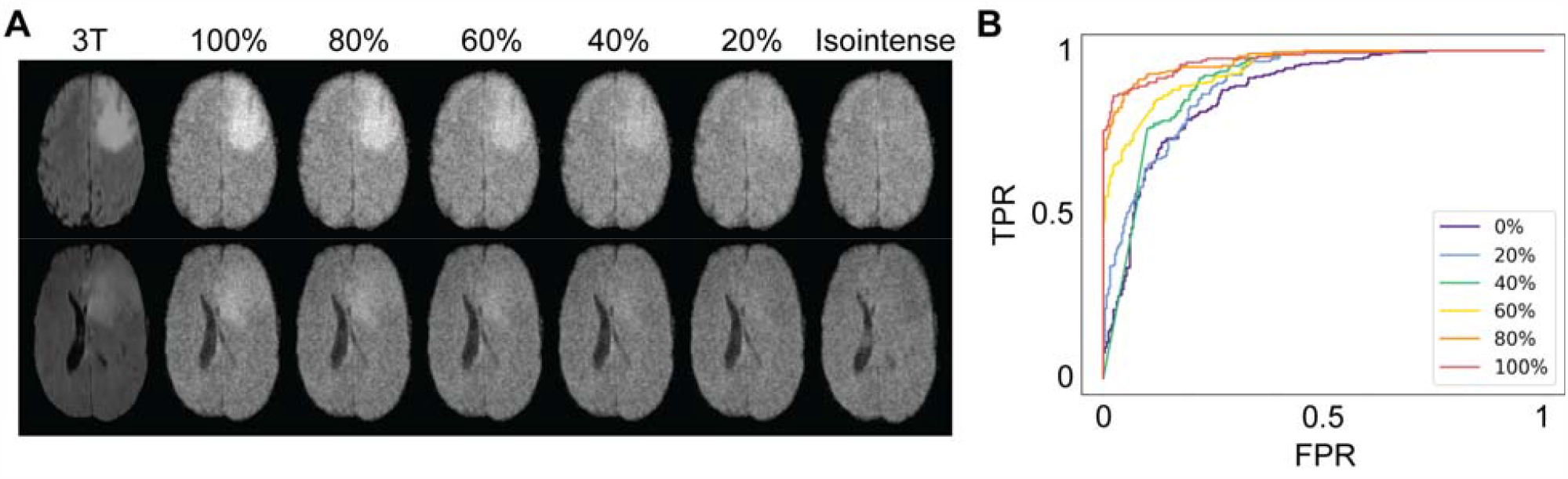
Intensity modulation to explore effects of intensity contrast. **(A)** Intensity values of the pathology segmentation were modulated over a range from normal intensity (100%) to isointense (tumor = background). **(B)** For each image subset, a classifier was trained to distinguish pathological and normal slices. AUC varied directly with lesion contrast but remained significantly better than chance even in the isointense cohort, which likely reflects structural deformations (such as ventricular effacement and midline shift as in the bottom row of panel A) or residual signal intensity heterogeneity.

## 4. Discussion

In this study we propose a generalizable methodology for evaluating novel imaging modalities, applied here to ULF MRI. By leveraging open-access datasets, this virtual trial paradigm permits rapid, low-cost assessment of a device’s potential diagnostic capabilities across a range of pathologies. We assert that this approach can help to address challenges in medical device development, regulatory approval, and clinical trial design.

ULF MRI offers an exciting opportunity for improving imaging accessibility in low-resource environments and enables point-of-care MRI. These scanners have relatively low manufacturing and operating costs and may help stem the increasing contribution of medical imaging to healthcare expenditures [33]. To accelerate device development cycles and reduce the cost of bringing devices to market, it is pivotal that tools are developed to allow rapid prototyping, efficient regulatory approval, and expedited deployment. Virtual clinical trials can efficiently evaluate medical imaging technology by simulating patients, imaging systems, and image interpreters [34]. For example, in breast tomography, Barufaldi et al. developed a virtual breast phantom and analytical pipeline that can simulate a clinical trial for several hundred patients per day [35]. While not a replacement for traditional prospective studies, simulated clinical trials may offer significant value as the FDA considers devices prior to extensive prospective data collection. Simulated trials could contribute to early feasibility studies (EFS) or provide supporting evidence for investigational device exemption (IDE) approval. Additionally, simulated trials could identify key patient populations or indications to prioritize for standard clinical trials. When considering a particular disease process, a simulated trial approach could help to establish benchmarks that a proposed device (such as a scanner or pulse sequence) must meet to provide sufficient diagnostic performance.

Based on simulated ULF images, our study suggests ULF MRI scanners should detect many brain lesions with comparable performance to standard MR imaging. However, simulated ULF imaging was sensitive to lesion size. Accuracy was lower for 1-4 cm^2^ lesions (approximately 2.25 cm diameter and below) as shown in **Fig. 4A**. These findings indicate that ULF MRI may perform adequately for identifying macro-scale pathology (most gliomas, medium-large vessel stroke, etc.) or measuring major brain structures, but may be less reliable for more subtle pathologies (MS lesions, embolic infarcts).

Importantly, we expect ULF MRI to complement and not replace standard clinical MRI. For this reason and as a proof of concept, our analysis is limited to basic diagnostic capabilities (presence vs absence of pathology), defining a range of expected size and signal intensity thresholds, rather than more complex image interpretation such as precise lesion segmentation or tracking lesion evolution over time. While we compare performance of ULF MRI to 3T MR devices, the practical alternative in certain use cases (ICU patients, underserved communities, in-office disease tracking) would be portable CT scans or no imaging at all. In these settings, ULF MRI may have advantages over CT such as increased tissue contrast and lack of ionizing radiation exposure.

This work underscores the power of open-access clinical databases to facilitate translational research. Platforms for data sharing, such as XNAT Central [17], iEEG Portal [36], and crowdsourced competitions [37] have led to rapid advances in many fields. While public databases provide diverse repositories of patient data with sufficient sample size to train deep learning algorithms, most medical imaging data remains federated across institutions [38]. Further data-sharing efforts designed explicitly for evaluating devices and software for regulatory approval could reduce the cost and time necessary to bring innovative imaging technology to the clinic. Recently radiology has shifted toward centralizing algorithms while maintaining individual data ownership [39]. While this approach may facilitate algorithm validation across research groups, it precludes creative use of multi-institutional data for applications beyond algorithm testing.

The simulated trial paradigm presented here is meant to serve as a framework for applying pre-existing datasets and deep learning to explore the expected performance of novel diagnostic devices. However, the approach brings with it several important limitations and methodological considerations. Most importantly, the utility of simulated data is directly linked to the transformation method quality. Here, we implemented a relatively simple histogram matching based algorithm. While the present method approximates tissue intensity and resolution in ULF images, it does not account for other potential field strength, pulse sequence and device-specific artifacts that may affect lesion conspicuity and image quality. More advanced transformation methods, including generative adversarial networks (GANs), synthetic MRI, and other quantitative methods [40–43] could certainly improve simulation quality. Future investigations should incorporate more advanced transformation methods and image quality validation by expert radiologist review.

However, transformation algorithms that learn by example, such as GANs, require large amounts of data. Methods that simulate images with a low N can be advantageous in certain situations [42]. Specifically, low-data requirement methods can be useful for evaluating new devices or during the prototyping process, where available data are scarce. While data-driven methods may produce closer matched simulations in the long run, these methods may not be applicable for all applications. Importantly, the simulated trial approach is not meant to serve as a replacement for prospective clinical trials. While more advanced methods such as GANs may improve image-to-image simulation quality, there will still likely be a gap between real and simulated images, especially in patients with pathology [43]. Simulations can provide useful guidance, including expected outcomes for prospective trials, however these are retrospective analyses and cannot provide the same level of scientific evidence as clinical trials.

Another consideration is the specific deep learning algorithm implemented. The purpose of the algorithm in this study was to provide a standardized baseline for analysis and comparison, and not necessarily to optimize detection for any particular use case. Approaches such as data augmentation or incorporation of data from multiple adjacent slices could likely provide incremental improvements in performance. Additionally, this work is limited by the sequences and pathologies that are publicly available. Results are likely sequence dependent and patients included in these datasets may not be reflective of the disease more broadly. Publicly available patient data is often collected at tertiary care centers and may be more likely to contain advanced pathology or larger lesions.

## 5. Conclusion

In this study, we have proposed a method for evaluating imaging devices via simulated trials, incorporating domain transfer and automated pathology detection, and demonstrated its application to a new ULF MRI device. This method allows for rapid evaluation of actual or proposed diagnostic devices. Importantly, simulated trials can provide guidance and justification for prospective studies. In our simulations, we found that gliomas, strokes and multiple sclerosis could be detected in ultra-low-field quality images. These findings justify prospective studies evaluating these pathologies on the device. This work additionally highlights the importance of centralized data sharing for device design and validation.

## Supporting information

STARD checklist

## Data Availability

All code related to simulated image generation, classifier design, and statistical analysis can be found at: https://github.com/tcama/SCT and https://github.com/penn-cnt/Arnold_simulated_clinical_trial.

https://www.oasis-brains.org/

http://www.ia.unc.edu/MSseg/

https://www.med.upenn.edu/cbica/brats2019.html

http://www.isles-challenge.org/ISLES2015/

https://portal.fli-iam.irisa.fr/english-msseg/

## Acknowledgment

We thank Jonathan Rothberg, PhD, and the team at Hyperfine Research (Guilford, GT), particularly Samantha By, PhD and Brian Welch, PhD, for the use of the ULF MRI scanner. We also thank the Penn Neuroradiology Research Core for assisting with patient recruitment and scanning. This work was supported by a sponsored research agreement with Hyperfine. Additional funding was provided by the NIH (T32NS091006-01), the HHMI-NIBIB Interfaces Initiative (5T32EB009384-10), Jonathan and Bonnie Rothberg, The Mirowski Family Fund, and Neil and Barbara Smit.

## CRediT Author Contributions

**T. Campbell Arnold**: Conceptualization, Methodology, Software, Formal Analysis, Writing – Original Draft, Writing – Review & Editing. **Steven N. Baldassano**: Conceptualization, Methodology, Software, Formal Analysis, Writing – Original Draft, Writing – Review & Editing. **Brian Litt**: Conceptualization, Writing – Review & Editing, Supervision, Funding acquisition. **Joel M. Stein**: Conceptualization, Writing – Review & Editing, Supervision, Funding acquisition.

## Abbreviations

(ULF): Ultra-low-field strength
(HF): high-field strength
(LGG): low-grade glioma
(HGG): high-grade glioma
(MS): multiple sclerosis
(FLAIR): fluid-attenuated inversion recovery
(SNR): signal to noise ratio
(AUC): area under curve
(ROC): receiver operating characteristic
(CAMs): class activation maps
(EFS): early feasibility studies
(IDE): investigational device exemption

