## Supplemental material for "Simulated Diagnostic Performance of Ultra-Low-Field MRI: Harnessing Open-Access Datasets to Evaluate Novel Devices"

**Supplemental Materials**

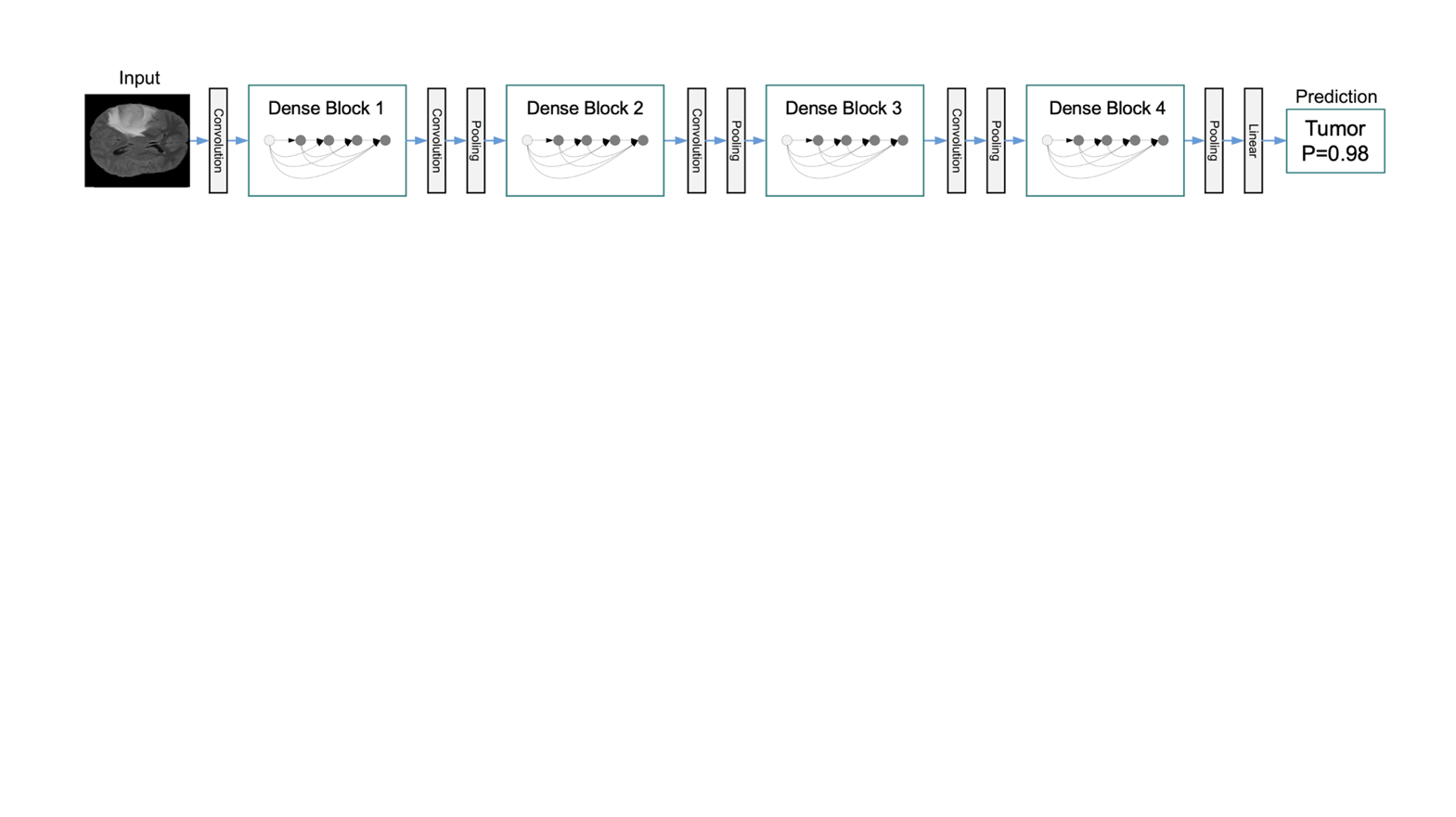

**Supplemental Figure 1:** Model architecture. Input to the model is a single axial MR image (224 x 224 pixels). The model consists of 4 dense blocks separated by transitional blocks containing convolutional and pooling layers. Details of the computational blocks can be found in the original paper by Huang, et al., at <https://arxiv.org/abs/1608.06993>. The output is a binary classification (lesion versus no lesion) with a prediction probability.

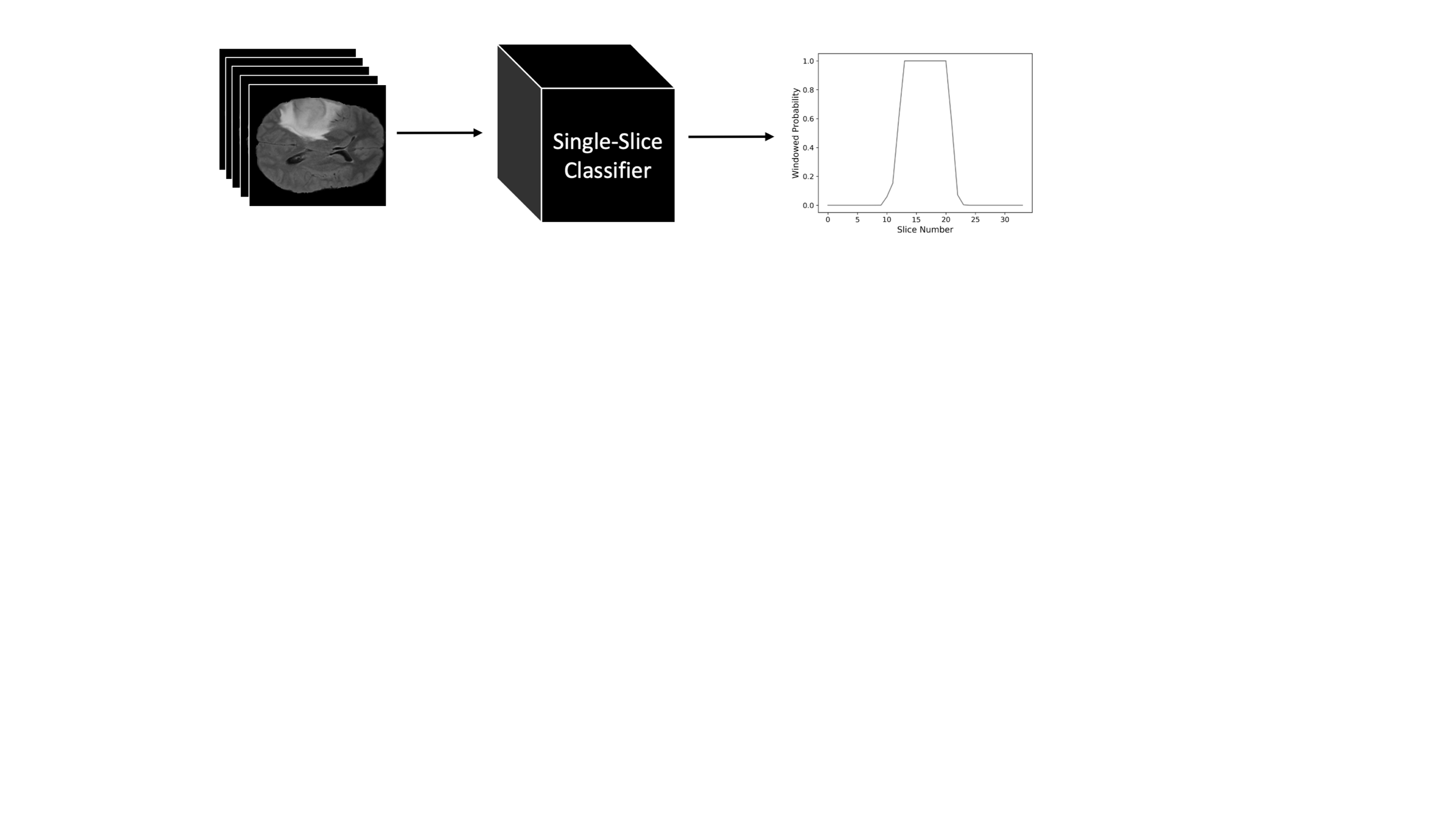

**Supplemental Figure 2:** Subject-level classification. The subject-level classifier builds on the single-slice classifier. Each axial slice in a subject’s image is classified using the single-slice classifier to generate a predicted lesion probability. A convolutional filter (incorporating approximately 1.5cm) is passed over the contiguous prediction probabilities to generate a locally windowed average. If the convolutional filter outputs a probability above a specified threshold, then the subject is classified as containing pathology.

| **Dataset** | **Num Training Images** | **Num Validation Images** |
| --- | --- | --- |
| LGG, 3T | 10,230 | 1,085 |
| LGG, low-field | 2,376 | 252 |
| HGG, 3T | 35,883 | 3,797 |
| HGG, low-field | 8,334 | 882 |
| MS, 3T | 10,824 | 1,428 |
| MS, low-field | 812 | 124 |
| Stroke, 3T | 2,553 | 614 |
| Stroke, low-field | 865 | 143 |

**Supplemental Table 1:** Training and validation split. Each dataset was split into training and validation sets with an approximate 9:1 division. A total of 8 classifiers were trained (4 pathologies, with separate 3T and simulated low-field classifiers). Abbreviations: High Grade Glioma (HGG), Low Grade Glioma (LGG), Multiple-Sclerosis (MS).
